# Landscape of copy number variants in Spanish People with Dementia

**DOI:** 10.64898/2026.01.26.26344703

**Authors:** Itziar de Rojas, Pablo García-González, Clàudia Olivé, Raquel Puerta, Laura Montrreal, Montserrat Alegret, Oscar Sotolongo-Grau, Amanda Cano, Marta Marquié, Sergi Valero, Miguel Calero, Alberto Rábano, Ana Belén Pastor, Teodoro del Ser, Inés Quintela, Juan Macías, Anaïs Corma-Gómez, Juan A Pineda, Emilio Franco-Macías, Dolores Buiza-Rueda, María Bernal Sánchez-Arjona, Jose Luis Royo, Silvia Mendoza, Carmen Lage, Carmen Antúnez, Arturo Corbatón-Anchuelo, María Teresa Martínez-Larrad, Mónica Diez-Fairen, Ignacio Alvarez, Raquel Huerto Vilas, Alfonso Arias Pastor, Manuel Menéndez-González, Carmen Martínez Rodríguez, Irene Rosas Allende, Sebastián García-Madrona, Ana Frank-García, Angel Martín-Montes, Miquel Baquero, GR@ACE, DEGESCO, Jordi Pérez-Tur, María J. Bullido, Guillermo Garcia-Ribas, Victoria Álvarez, Gerard Piñol-Ripoll, Pau Pastor, Eloy Rodriguez-Rodriguez, Jose María García-Alberca, Pablo Mir, Luis M Real, Miguel Medina, María Eugenia Sáez, Ángel Carracedo, Michael T. Heneka, Lluís Tàrraga, Mercè Boada, Pascual Sánchez-Juan, M. Victoria Fernández, Sven J. van der Lee, Agustín Ruiz

**Affiliations:** Ace Alzheimer Center Barcelona, Universitat Internacional de Catalunya, Barcelona, Spain.; CIBERNED, Network Center for Biomedical Research in Neurodegenerative Diseases, National Institute of Health Carlos III, Madrid, Spain; Luxembourg Centre for Systems Biomedicine (LCSB), University of Luxembourg, Esch-sur-Alzette, Luxembourg; Doctorate in Biotecnology, Faculty of Pharmacy and Food Sciences, University of Barcelona, Barcelona, Spain.; CIEN Foundation/Queen Sofia Foundation Alzheimer Center/Instituto de Salud Carlos III; UFIEC, Instituto de Salud Carlos III; CIEN Foundation/Queen Sofia Foundation Alzheimer Center; BT-CIEN; Department of Neurology/CIEN Foundation/Queen Sofia Foundation Alzheimer Center; Grupo de Medicina Xenómica, Fundación Pública Galega de Medicina Xenómica, Santiago de Compostela, Spain.; Instituto de Biomedicina de Sevilla (IBIS)/Departamento de Medicina, Universidad de Sevilla, Sevilla. Spain; CIBERINFEC, Network Center for Biomedical Research in Infectious Diseases, National Institute of Health Carlos III, Madrid, Spain; Unidad Clínica de Enfermedades Infecciosas y Microbiología. Hospital Universitario de Valme, Sevilla, Spain; Unidad de Demencias, Servicio de Neurología y Neurofisiología. Instituto de Biomedicina de Sevilla (IBiS), Hospital Universitario Virgen del Rocío/CSIC/Universidad de Sevilla, Seville, Spain; Unidad de Trastornos del Movimiento, Servicio de Neurología y Neurofisiología. Instituto de Biomedicina de Sevilla (IBiS), Hospital Universitario Virgen del Rocío/CSIC/Universidad de Sevilla, Seville, Spain; Departamento de Especialidades Quirúrgicas, Bioquímica e Inmunología. School of Medicine. University of Malaga. Málaga, Spain; Alzheimer Research Center & Memory Clinic, Instituto Andaluz de Neurociencia, Málaga, Spain.; Neurology Service, Marqués de Valdecilla University Hospital (University of Cantabria and IDIVAL), Santander, Spain.; Unidad de Demencias, Hospital Clínico Universitario Virgen de la Arrixaca, Murcia, Spain.; Instituto de Investigación Sanitaria, Hospital Clínico San Carlos (IdISSC), Madrid, Spain; Spanish Biomedical Research Centre in Diabetes and Associated Metabolic Disorders(CIBERDEM), Madrid, Spain; Fundació Docència i Recerca MútuaTerrassa, Terrassa, Barcelona, Spain; Memory Disorders Unit, Department of Neurology, Hospital Universitari Mutua de Terrassa, Terrassa, Barcelona, Spain; Unitat Trastorns Cognitius, Hospital Universitari Santa Maria de Lleida, Lleida, Spain; Institut de Recerca Biomedica de Lleida (IRBLLeida), Lleida, Spain; Servicio de Neurología. Hospital Universitario Central de Asturias, Oviedo, Spain; Instituto de Investigación Sanitaria del Principado de Asturias (ISPA); Departamento de Medicina, Universidad de Oviedo, Oviedo, Spain; Hospital de Cabueñes, Gijón, Spain; Laboratorio de Genética. Hospital Universitario Central de Asturias, Oviedo, Spain; Hospital Universitario Ramon y Cajal, IRYCIS, Madrid; None; Department of Neurology, La Paz University Hospital. Instituto de Investigación Sanitaria del Hospital Universitario La Paz. IdiPAZ.; Hospital La Paz Institute for Health Research, IdiPAZ, Madrid, Spain; Universidad Autónoma de Madrid; Department of Neurology, La Paz University Hospital; Servei de Neurologia, Hospital Universitari i Politècnic La Fe, Valencia, Spain; Grupo de Investigación en Enfermedad de Alzheimer, GINEA, Instituto de Investigación Sanitaria La Fe; Unitat de Genètica Molecular, Institut de Biomedicina de València-CSIC, Valencia, Spain; Centro de Biología Molecular Severo Ochoa (UAM-CSIC); Instituto de Investigacion Sanitaria ‘Hospital la Paz’ (IdIPaz), Madrid, Spain; Unit of Neurodegenerative diseases, Department of Neurology, Hospital Germans Trias i Pujol, Badalona, Barcelona; Neurodegenerative Diseases Research Laboratory, Germans Trias i Pujol Research Laboratory, Badalona, Barcelona; Unidad de Trastornos del Movimiento, Servicio de Neurología, Instituto de Biomedicina de Sevilla, Hospital Universitario Virgen del Rocío/CSIC/Universidad de Sevilla, Seville, Spain; Centro de Investigación Biomédica en Red sobre Enfermedades Neurodegenerativas, Instituto de Salud Carlos III, Madrid, Spain; Departamento de Medicina, Facultad de Medicina, Universidad de Sevilla, Seville, Spain; Department of Medical Biochemistry, Molecular Biology and Immunology, University of Sevilla, Sevilla, Spain; CAEBI, Centro Andaluz de Estudios Bioinformáticos, Sevilla, Spain; Grupo de Medicina Xenómica, CIBERER, CIMUS. Universidade de Santiago de Compostela, Santiago de Compostela, Spain.; Fundación Pública Galega de Medicina Xenómica- IDIS, Santiago de Compostela, Spain.; Alzheimer’s Centre Reina Sofia-CIEN Foundation-ISCIII, Madrid, Spain; Genomics of Neurodegenerative Diseases and Aging, Human Genetics, Vrije Universiteit Amsterdam, Amsterdam UMC location VUmc, Amsterdam, The Netherlands.; Alzheimer Center Amsterdam, Neurology, Vrije Universiteit Amsterdam, Amsterdam UMC location VUmc, Amsterdam, The Netherlands; Amsterdam Neuroscience, Neurodegeneration, Amsterdam, The Netherlands; Biggs Institute for Alzheimer’s and Neurodegenerative Diseases, University of Texas Health Science Center, San Antonio, Texas, USA

**Keywords:** Copy number variant, Structural variation, Neurogenetics, Alzheimer’s disease

## Abstract

**Background:** Recent studies suggest that copy number variants (CNVs) may contribute to the missing heritability of complex diseases such as Alzheimer’s disease (AD) and related dementias (ADRD).

**Methods:** We performed a CNV analysis using genotyping data (Axiom 815K Spanish biobank array) from the GR@ACE/DEGESCO dementia dataset (n=20,067) of the Spanish population. Applying PennCNV and extensive quality control, 8,275 controls and 7,818 dementia cases were selected for gene-level case/control associations.

**Results:** We identified 43,833 CNVs with deletions (47%) and duplications (53%). No genome-wide significant associations were found, but nominal associations were observed in *PKP3*-*SIGIRR* and *FBRSL1* loci. CNVs in 2,970 genes were exclusive to dementia cases, enriched in vascular-related pathways. Notable findings included 14q11.2 duplication and *VPS13B* deletions in ADRD cases, the latter confirmed by optical genome mapping.

**Conclusion:** Our findings suggest potential novel genes associated with ADRD in the Spanish population. However, the limited resolution of array-based technologies in detecting CNVs warrants further investigation.

## Background

Dementia is a clinical syndrome resulting from a variety of underlying conditions—both neurodegenerative and non-neurodegenerative—characterized by progressive cognitive decline and loss of functional independence. It poses a major global health challenge, affecting millions of people worldwide^1^. Genetics plays a critical role in the risk landscape of Alzheimer’s disease and related dementias (ADRD), as heritability estimates suggest that approximately 70–80% of disease susceptibility can be attributed to genetic factors^2^. Consequently, numerous genome-wide association studies (GWAS) and sequencing efforts have identified common and rare variants associated with these ADRD, substantially advancing our understanding of their genetic architecture ^3–5^. However, despite these advances, a significant proportion of heritability remains unexplained, suggesting additional genomic mechanisms at play. Among these, copy number variations (CNVs) remain relatively underexplored in ADRD^6,7^, leaving important gaps in our understanding of their role in neurodegeneration.

CNVs are one of the most prevalent types of structural variations (SV) in the human genome involving gain (duplication) or loss (deletion) of specific segments of DNA that vary from one kilobase (kb) to several megabases (Mb)^8^. Due to their large size, many CNVs span or disrupt multiple genes, resulting in altered gene dosage and impaired biological function^9^. This mechanism can underlie not only isolated gene disorders but also the development of contiguous gene syndromes or CNV syndromes (CGSs) caused by deletions or duplications that affect several neighboring genes^10^. Some CGSs, such as 22q11.2 deletion syndrome, 1q21.1 and 16p11.2 CNV syndromes, and Williams–Beuren syndrome (7q11.23 deletion) present with cognitive impairment, developmental delay, or even progressive neuropsychiatric symptoms^11^ that overlap with dementia phenotypes.

Importantly, several CNVs have been linked to several complex disorders including neurodegenerative diseases^9^ such as amyotrophic lateral sclerosis (e.g., *C9orf72* repeat expansions)^12,13^, Parkinson’s disease (*SNCA* duplications)^14,15^ and Alzheimer’s disease (AD; *APP* locus duplications)^8,16–20^. Beyond neurodegeneration, CNVs also increase risk for psychiatric and neurodevelopmental conditions such as schizophrenia^21^, autism, and intellectual disability^22,23^. These genomic rearrangements can alter gene expression, disrupt critical biological pathways involved in neuronal function, synaptic plasticity, immune response, and protein homeostasis, ultimately contributing to the onset and progression of dementia symptoms.

Recent advances in genomic technologies now allow systematic investigation of CNVs across large cohorts, revealing that up to 12% of the human genome and thousands of genes are variable in copy number responsible for an important proportion of normal phenotypic variation^24,25^. However, CNV detection in next-generation sequencing (NGS) data requires high-sequencing depth and uniformity of coverage, which may not be achievable in a cost- and time-effective manner^26,27^. Consequently, microarray-based CNV detection and its calling algorithms continues to be commonly used clinical genetic test^28^, offering an optimal compromise between cost and resolution and enabling the reuse of extensive existing datasets for research.

Current challenges include linking CNVs to biological function and understanding their role in common and complex human diseases. Large-scale GWAS and integrative multi-omics approaches are now essential to identify novel CNVs associated with dementia and decipher their functional consequences^29,30^. Addressing these gaps could advance personalized medicine strategies and guide the development of targeted therapies.

Here, we investigated the genome-wide landscape of CNVs and their associations with ADRD in a large cohort of 20,067 participants from the Spanish GR@ACE/DEGESCO consortium in order to further characterize the genetic causes of dementia in this population. We report CNVs overlapping known genes such as *APP, MAPT* and *ABCA7*. Additionally, we also describe novel CNVs potentially associated with dementia, overlapping the genes *FBRSL1, PKP3-SIGIRR, VPS13B* and 14q11.2 region.

## Methods

### GR@ACE/DEGESCO samples

The individuals that participated in the study were obtained from multiple Spanish centers collected by GR@ACE/DEGESCO consortium. The GR@ACE cohort^31,32^ recruited dementia continuum patients from Ace Alzheimer Center Barcelona (Barcelona, Spain), and control individuals from three centers: Ace Alzheimer Center Barcelona, Valme University Hospital (Seville, Spain), and the Spanish National DNA Bank–Carlos III (University of Salamanca, Spain) (http://www.bancoadn.org). Additional dementia, mild cognitive impairment (MCI) cases and population healthy controls were obtained from 13 research centers included in Dementia Genetic Spanish Consortium (DEGESCO)^33^.

At all sites, AD diagnosis was established by a multidisciplinary working group—including neurologists, neuropsychologists, and social workers—according to the DSM-IV criteria for dementia and the National Institute on Aging and Alzheimer’s Association’s (NIA–AA) 2011 guidelines for diagnosing AD. For MCI diagnosis, the classification of López et al.^34^ and Petersen’s criteria were also used^35–37^. For further details ^5,31,32^ on the contribution of the sites, see Supplementary Table 1. Written informed consent was obtained from all the participants. The ethics and scientific committees have approved this research protocol (Acta 25/2016, Ethics Committee H., Clinic I Provincial, Barcelona, Spain).

Methods for genetic data acquisition and detailed quality control (QC) procedures have been previously described^31,32^. Genotyping was conducted using the Axiom 815K Spanish biobank array (Thermo Fisher) at the Spanish National Center for Genotyping (CeGEN, Santiago de Compostela, Spain). SNP-arrays are used to evaluate the intensity of the hybridization signal of genomic DNA with the average value of control DNA using single-source hybridization instead of competitive hybridization, being able to detect submicroscopic CNVs^38^.

### CNV detection and quality control

CNV calls were generated using PennCNV^39^ v.1.0.5 on raw signal intensity data from each genotyping array. The PennCNV-Affy protocol (http://www.openbioinformatics.org/penncnv/penncnv_tutorial_affy_gw6.html) was first performed to transform raw CEL files into a signal intensity file containing the Log R Ratio (LRR) as a normalized measure of the total signal intensity for two alleles of the SNP and, the B Allele Frequency (BAF) which is a normalized measure of the allelic intensity ratio of two alleles to generate CNV calls. We followed the user guidelines of PennCNV^39^ and used the Hidden Markov Model^39,40^ ‘‘affygw6.hmm’’ and a population frequency of B allele (PFB file) calculated with the coordinates from the Axiom SpainBA array. The combination of LRR and BAF was used to determine the presence of CNVs in the genome^41,42^ and we extracted all duplications and deletions covering the genome (HG build 37).

Extensive QC of CNV calls was performed on all samples, restricting the analyses to autosomes (Supplementary Table 2). Adjacent CNVs were merged where the gap was <30% of total length (--fraction 0.3). Quality metrics included CNV call counts, LRR standard deviation (SD), BAF Drift and Waviness Factor (WF). Samples were excluded if they had an excessive number of CNVs (>50 CNV) and exceed any of the following thresholds: LRR SD>0.38, BAF Drift>0.015 or WF>0.1. To ensure we were including only high-confidence CNVs in the analysis, CNVs called based on data <10 SNPs, length <50KB and CNVs that had >50% overlap with chromosomal centromeric and telomeric, or immunoglobulin regions as defined in Need et al.^43^ plus 250kb were excluded as they are known to be prone to artificial CNV calls and therefore false discoveries.

Individuals with low-quality SNP raw data, excess of heterozygosity, sex discrepancies and familial relations between samples (PI-HAT > 0.1875) were excluded from the analysis. A principal component analysis (PCA) was performed, and population outliers were removed (European cluster of 1000 Genomes). Additionally, individuals with Down Syndrome were excluded from the analyses due to trisomy chr21 and the high probability of structural DNA alterations.

CNVs may involve one or more genes and are distributed in a non-uniform manner. In this sense, we annotated the overlapping genes (partially or totally) by each CNV using the reference gene annotation (refGene.txt.gz and refLink.txt.gz) from UCSC Genome Browser build 37. To minimize potential batch effects related to CNV calling, we performed an additional PCA based on the presence/absence of annotated gene deletions and duplications. Individuals falling outside ±3 SD from the mean of PC1 were excluded as potential outliers. Subsequently, individuals carrying any of the 15 genes indicated by Aguirre et al.^29^ based on data from UK Biobank, classified as genes with greater intolerance to structural variation (*BRCA2, BRCA1, APC, ATM, MSH2, MLH1, MSH6, RB1, SBDS, SPATA31D1, CYP3A4, PABPC3, OTOP1, KRT16* and *ZNF302*), were excluded. These genes were enriched in early-onset diseases, particularly cancer^29,44–46^. Afterwards, to avoid batch effects, analysis of variance (ANOVA) comparisons between ctrl-ctrl individuals based on different sources of DNA origin (i.e. coming from extracted DNA, blood, saliva or DNA banks) were performed in duplications and deletions separately by removing genes with p-value<0.001. Finally, 16,093 samples (7,818 dementia cases and 8,275 controls) with 43,833 CNV calls remained after all QC measures and were entered into case/control association analyses (Supplementary Table 2).

Comparisons between dementia cases and controls were performed using the Wilcoxon rank-sum test for continuous variables (e.g., CNV count, length, genes), and chi-square tests for categorical comparisons (CNV presence and sex distribution). All analyses were conducted in R (version 4.4.1), p-values < 0.05 were considered statistically significant.

### CNV association analyses

Gene-level dementia case/control association tests were conducted across all genes in a 16,093 unrelated individuals of Spanish ancestry using linear regression implemented in Plink2^47^ (www.cog-genomics.org/plink/2.0/) including the 4PCs based on the annotated gene CNVs as covariates. Gene burden was encoded as a binary variable which indicates whether an individual has a CNV which contains any overlap of the HUGO Gene Nomenclature Committee (HGNC) gene region. For CNVs spanning multiple genes, each affected gene was individually considered in the analysis. We treat deletions and duplications identically, with the assumption that any CNV which overlaps a gene in this fashion will disrupt its normal function. 3,035 genes were considered in the genome-wide approach for deletions, and 5,192 for duplications. Bonferroni for multiple testing correction was applied (1.65×10^-05^ and 9.6×10^-06^ respectively). Another case-only analysis was performed to identify genes that may play a role in dementia susceptibility using Fisher Test.

The quantity and quality of CNVs reported by different algorithms and different platforms exhibit substantial variance, therefore, it is often recommended to visually examine CNV calls to judge whether they are reliable or not. Candidate CNVs were visually inspected with R software (4.4.2) using the BAF al LRR values.

### CNV burden analyses

To determine whether dementia cases show a greater number of CNVs (deletions and duplications) t-test were performed. Furthermore, burden analysis (Fisher Test) of AD genes affected by CNVs were conducted between dementia cases and controls, including 76 AD genes identified from the latest meta-GWAS by the European Alzheimer’s and Dementia consortium^5^.

### Optical genome mapping

Optical genome mapping (OGM)^48,49^ is a high-resolution genomic analysis technique that offers a comprehensive view of an individual’s DNA structure. This cutting-edge technology uses advanced imaging of very long linear single DNA molecules (median size > 250 kb) and computational methods to map their arrangement within the genome. Thus, OGM was used for an independent experimental validation of CNVs in the *VPS13B* gene regions of the individuals for whom we had blood availability (n=3 out of 9 samples).

#### DNA extraction, labeling, and chip loading

For each sample, ultra-high molecular weight (UHMW) genomic DNA (gDNA) was extracted from 650μL of frozen blood following manufacturer’s protocols (Bionano Prep SP Blood and Cell DNA Isolation Kit; Bionano Genomics, San Diego, CA). Briefly, after thawing in 37 °C water bath, white blood cells were counted, pelleted (2200g for 2min), and treated with proteinase K (Qiagen) and LBB lysis buffer to release gDNA. After phenylmethylsulfonyl fluoride treatment (PMSF; Sigma-Aldrich, St. Louis, MO) to inactivate proteinase K, a Nanobind paramagnetic disk was added to the solution, mixed with isopropanol to precipitate the gDNA and washed with WB1 and WB2. After transferring to a new tube, buffer EB (Bionano Genomics, San Diego, CA) was added to elute the gDNA from the disk. The gDNA was equilibrated overnight at room temperature for homogenization and subsequently quantified with Qubit Fluorometer (Invitrogen™ Qubit™ 4 Fluorometer).

Sequence-specific Direct label and stain (DLS) technique was used for labeling following manufacturer’s protocols (Bionano Prep DLS Labeling Kit; Bionano Genomics, San Diego, CA). Direct Labeling Enzyme 1 (DLE-1) reaction was carried out using 750 ng of gDNA to tag a specific 6 bp sequence (CTTAAG) with a DL-green fluorophore (∼15 times per 100 kb). Following Puregene proteinase K (Qiagen, Hilden, Germany) digestion and DL-Green clean-up using DLS membrane in DLS 24-well plate, the labeled gDNA was mixed with DNA Stain, stained overnight at room temperature for backbone visualization and quantified using Invitrogen™ Qubit™ 4 Fluorometer. The fluorescent-labeled gDNA molecules (4-12ng/µl) were loaded on a Saphyr chip G2.3, and linear double stranded molecules passing across nanochannels were sequentially imaged on a Saphyr instrument (Bionano Genomics, San Diego, CA).

#### Data analysis and variant filtering

Effective genome coverage of at least 160X effective coverage was achieved for *de novo* assemblies. Standard run quality control parameters [total DNA (≥150 kbp), N50 (≥150 kbp), map rate (≥150 kbp), effective coverage (>300x), average label density (per 100 kbp)] were evaluated per manufacturer’s guidelines. Details regarding limit of detection, reproducibility and precision are described elsewhere^50,51^.

The *de novo* assembly (DN; SVs > 500 bp) pipeline was executed on Bionano Solve software v3.7. (Bionano Genomics, San Diego, CA). Reporting and direct visualization of structural variants was done on Bionano Access v1.7.1. using Genome Reference Consortium GRCh38/hg38 as the reference.

SVs were identified based on discrepancies in the labeling pattern between the sample and the reference. The complementary coverage based CNV algorithm identified large chromosomal gains and losses (˃500kbp). Confidence scores were calculated and provided by Bionano Genomics^50^. For data filtering, the following recommended confidence scores were used: Insertion/Deletion=0; Inversion=0.7; Duplication=-1; Intra-fusion/Inter-translocation=0.05; Copy Number=0.99; Aneuploidy=0.95.

Genome imaging generated at least 550 Gb of data per sample later analyzed with the DN pipeline. Label density was 14.24 labels/100 kb on average. The average N50 molecule length (molecules ≥150 kb and at least 9 labels) was 272.63 kb. Only molecules over 150 kb were included. Tall of this resulted in an average map rate of 79% and an effective coverage of 196.8X for DN pipeline (Supplementary Table 3).

### Post-hoc analyses

In the human genome, CNVs are also classified into several categories: benign CNV (normal genomic variant), likely benign CNV, variant of uncertain significance, CNV of possible clinical relevance (high-susceptibility locus/risk factor/likely pathogenic variant), and clinically relevant CNV (pathogenic variant). Information on the *VPS13B* gene region was extracted from ClinVar^52,53^ and comparisons were conducted to determine whether the identified CNVs had been previously described or appeared to be novel.

Regarding SNPs, all those reported in ClinVar with high imputation quality (Rsq>0.3) were extracted from the array, and an AD/Ctrl association was performed to assess whether other types of mutations affected the *VPS13B* region. Additionally, a burden analysis was conducted to evaluate the cumulative effect. Individual-level variant counts were calculated by summing heterozygous and homozygous minor alleles across all markers. Generalized linear models (GLMs), including Poisson and negative binomial regressions (NBR), and non-parametric testing (Wilcoxon) were applied to test for differences in variant burden between AD cases and controls. Distribution of variant burden per individual was visualized using violin and density plots.

The Genome Aggregation Database SV resource^54^ (GnomAD CNVs v4.1.0) was used as a reference for the types and frequencies of CNVs in the genes of interest within a larger cohort (includes 464,297 unrelated exomes). In addition, gene transcript information was extracted from this database.

Furthermore, the ClassifyCNV^55^ software was employed for the accurate identification and clinical annotation of CNVs. This tool implements the 2019 ACMG classification guidelines to assess CNV pathogenicity using genomic coordinates and CNV type as input. It provides a clinical classification for each variant, along with a breakdown of the classification score and a list of genes potentially relevant for variant interpretation.

#### Pathway analysis

Pathway enrichment analysis was conducted in R (4.4.2) using the genes affected by CNVs observed exclusively in at least four dementia cases. Gene annotations were converted to Entrez IDs, and both KEGG and Gene Ontology (GO, Biological Process) enrichment analyses were performed using the clusterProfiler() package. Significance thresholds were set at p-value < 0.05 and q-value < 0.2, with multiple testing correction applied using the Benjamini-Hochberg method.

## Results

### Sample demographics and CNV call characteristics

We analyzed a total of 43,833 CNVs across 16,093 individuals, including 8,275 controls and 7,818 dementia cases. Dementia cases included a significantly higher proportion of females (68.6% vs. 50.0% in controls) and were substantially older, as expected (mean age: 79.9 vs. 56.9 years) compared to the population control group. However, the proportion of individuals carrying at least one CNV was comparable between groups (88.7% in controls vs. 88.0% in dementia, p = 0.208, Table 1). Overall, the detected CNV represented 19,579 unique genomic regions. Notably, only 2,380 (12.2%) CNV regions were shared between cases and controls, indicating that the majority of CNV regions were exclusive to either group, reflecting a high degree of non-overlap in CNV distribution. On average, control individuals carried more CNVs (2.81 vs 2.64, p = 3.2×10^-03^). As expected, CNV length and the number of SNPs within each CNV were strongly correlated overall (r = 0.82), supporting the reliability of probe density as a proxy for CNV size. Interestingly, this correlation was stronger among controls (r = 0.85) compared to dementia cases (r = 0.76). This may help explain why, although dementia cases having slightly longer CNVs on average (272.7 kb vs. 262.3 kb, p = 3.06×10^-02^), the average number of SNPs within CNVs was lower (71.0 vs. 78.9 SNPs, p = 3.4×10^-04^). In addition, regression analysis adjusted by sex and diagnosis showed that age was positively associated with CNV length (p = 1.2×10^-03^), while no significant association was observed with the number of SNPs per CNV (p = 0.21). These differences suggest that CNVs in dementia cases may more frequently affect genomic regions with lower SNP density, and that age-related increases in CNV size do not necessarily correspond to higher probe density. Nevertheless, the number of genes affected per CNV and per individual was largely comparable across groups (p>0.05, Table 1).

**Table 1:** Cohort demographic and CNV characteristics. *Chi-square test used for comparisons of sex and proportion of individuals with CNVs; Wilcoxon rank-sum test used for all other comparisons.

|  | *Control* | *Dementia* | *Total* | *P-value\** |
| --- | --- | --- | --- | --- |
| **Individuals (n,%)** | 8,275 (51.42) | 7,818 (48.58) | 16,093 | - |
| **Indiv. with CNVs (n,%)** | 7,339 (88.7) | 6,883 (88.0) | 14,222 (88.4) | 0.208 |
| **Females (n,%)** | 4,139 (50) | 5,362 (68.6) | 9,501 (59.0) | <2.2x10<sup>-16</sup> |
| **Age (mean, sd)** | 56.9 (17.9) | 79.9 (7.88) | 68.17 (18.06) | <2.2x10<sup>-16</sup> |
| **CNVs (n,%)** | 23,217 (52.97%) | 20,616 (47.03%) | 43,833 | - |
| **Unique** | 9,498 (48.5%) | 7,701 (39.3%) | 19,579 | - |
| **Per individual** | 2.81 | 2.64 | 2.72 | 3.16x10<sup>-03</sup> |
| **SNPs (mean, sd)** | 78.9 (436) | 71.0 (347) | 75.2 (397.0) | 3.42x10<sup>-04</sup> |
| **Length (mean kb, sd)** | 262.3 (1503.7) | 272.7 (1250.5) | 267.2 (1390.4) | 3.06x10<sup>-02</sup> |
| **Genes affected by CNV (mean, sd)** | 2.93 (5.1) | 2.94 (4.3) | 2.94 (4.7) | 0.989 |
| **Genes affected by indiv (mean, sd)** | 8.79 (34.6) | 8.12 (22.0) | 8.46 (29.18) | 0.079 |
| **Duplications (n,%)** | 11,963 (51.8%) | 11,115 (48.2%) | 23,078 | - |
| **Per individual** | 1.45 | 1.42 | 1.43 | 0.060 |
| **SNPs (mean, sd)** | 96.5 (537) | 87.5 (442) | 92.2 (493) | 1.42x10<sup>-09</sup> |
| **Length (mb)** |  |  |  |  |
| min-max | 0.050-146.10 | 0.050-133.62 | 0.050-146.10 | 0.299 |
| mean-sd | 0.340 (1.933) | 0.355 (1.640) | 0.347 (1.798) |  |
| **Genes affected by CNV (mean, sd)** | 3.91 (7.3) | 3.89 (10.5) | 3.9 (9.0) | 0.247 |
| **Genes affected by indiv (mean, sd)** | 5.93 (22.6) | 5.68 (19.4) | 5.81 (21.1) | 0.155 |
| **Deletions (n,%)** | 11254 (54.2%) | 9501 (45.8%) | 20,755 | - |
| **Per individual** | 1.36 | 1.22 | 1.29 | 0.041 |
| **SNPs (mean, sd)** | 60.1 (294) | 51.8 (181) | 56.3 (248) | 0.135 |
| **Length (mb)** |  |  |  |  |
| min-max | 0.050-74.722 | 0.050-29.353 | 0.50-74.722 | 0.758 |
| mean-sd | 0.180 (0.825) | 0.176 (0.477) | 0.178 (0.688) |  |
| **Genes affected by CNV (mean, sd)** | 2.20 (3.9) | 2.25 (3.3) | 2.2 (3.6) | 0.976 |
| **Genes affected by indiv (mean, sd)** | 3.62 (19.0) | 3.10 (12.4) | 3.37 (16.1) | 0.143 |

Comparing duplications vs. deletions, duplications were significantly more frequent than deletions (23,078 vs. 20,755 events, p < 2.2×10^-16^); were consistently larger, contained more SNPs, and affected more genes per CNV and per individual (mean CNVrate_del-dup_= 1.29-1.43, mean length_del-dup_= 178-347.4 kb and mean genes_del-dup_= 3.4-5.8; p < 2.2×10^-16^). When stratified by CNV type, duplications were slightly more frequent in controls (51.8%) than in dementia. On average, individuals carried ∼1.4 duplications. Duplication length was comparable (∼0.35 Mb), and they affected ∼3.9 genes per CNV and ∼5.8 genes per individual. Although gene burden did not differ significantly, duplications in individuals with dementia contained significantly fewer SNPs (p = 1.42×10^-09^). Deletions occurred significantly more frequently in controls (1.36 vs. 1.22; p = 0.041), though deletion length and gene content did not differ between groups (Table 1). Finally, linear regression models adjusted for age, sex and diagnosis showed no significant association between age or sex and the number of deletions or duplications per individual (p>0.05).

### Case/control association analyses

#### Genome-wide approach

We identified 7,177 of the 28,295 genes tested (25.4%) to be overlapped by CNV calls. We did not find genome-wide CNVs associated with dementia in this population, both in deletions and duplications using linear regression (Bonferroni-corrected p=1.65×10^-05^ and 9.63×10^-06^ respectively, Figure 1). However, we observed nominal deletions-associations with dementia at chr11, overlapping *PKP3* (median=93.9kb, p=1.05×10^-03^) and *SIGIRR* genes (median=98.7Kb, p=2.23×10^-03^), as well as at chr12 overlapping *FBRSL1* gene (median=97.2kb, p=2.18×10^-04^, Supplementary Figure 1). Although these associations did not exceed the multiple testing threshold, the regions identified may represent promising loci for follow-up in larger cohorts or functional studies.

**Figure 1:**
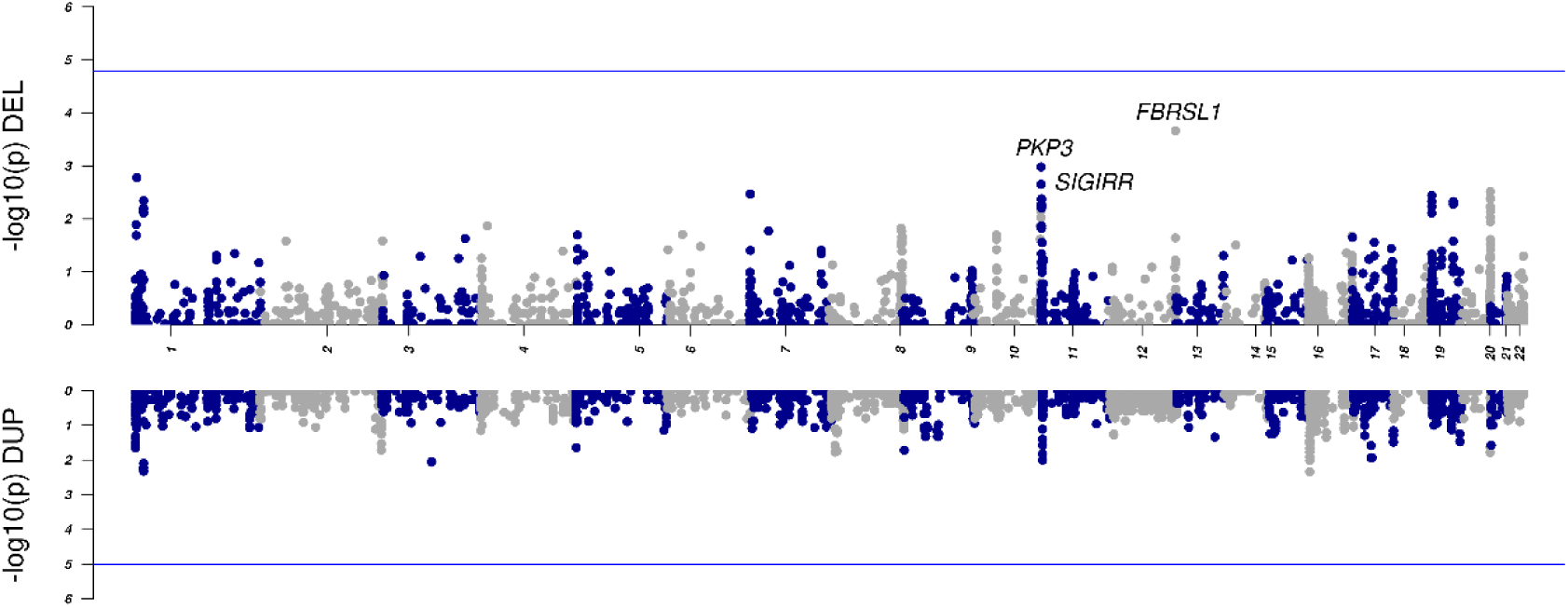
Miami plot for logistic regression in deletions (λ=0.661) and duplications (λ=0.347). Genome-wide threshold was set to p<1.65E-05 for deletions and p<9.63E-06 for duplications (Bonferroni correction).

### Case-only CNV findings

We identified 2,970 genes across the genome that were overlapped by CNV calls exclusively in dementia cases, with no overlaps observed in controls from the GR@ACE/DEGESCO study. Genes overlapped by CNV calls from at least four independent cases are shown in Supplementary Table 4, suggesting that may represent novel structural abnormalities with potential functional relevance to dementia pathogenesis. Pathway enrichment analysis of these genes highlighted significant involvement in cardiac muscle adaptation, hypertrophic signaling, and vascular response pathways, indicating that these CNVs may influence disease risk through mechanisms involving cardiovascular remodeling and dysregulation (Supplementary Table 5).

A duplication encompassing the 14q11.2 region was identified exclusively in AD patients (n=8) and was absent in controls (Supplementary Table 6). This duplication is located on chromosome 14 and fully spans the *CMTM5, EFS, IL25, MHRT, MIR208A, MIR208B, MYH6, MYH7* genes, extending both upstream and downstream in all cases, with an estimated size ranging from 78.7 to 146.1 Kb.

We also observed intragenic deletions affecting the Vacuolar Protein Sorting 13 Homolog B gene (*VPS13B*) in chromosome 8 in 9 dementia patients. Deletions occurs quite uniformly in all cases, affecting 82-254.8 Kb range and exons 18-33 out of 62 (Figure 2, Table 2).

**Figure 2:**
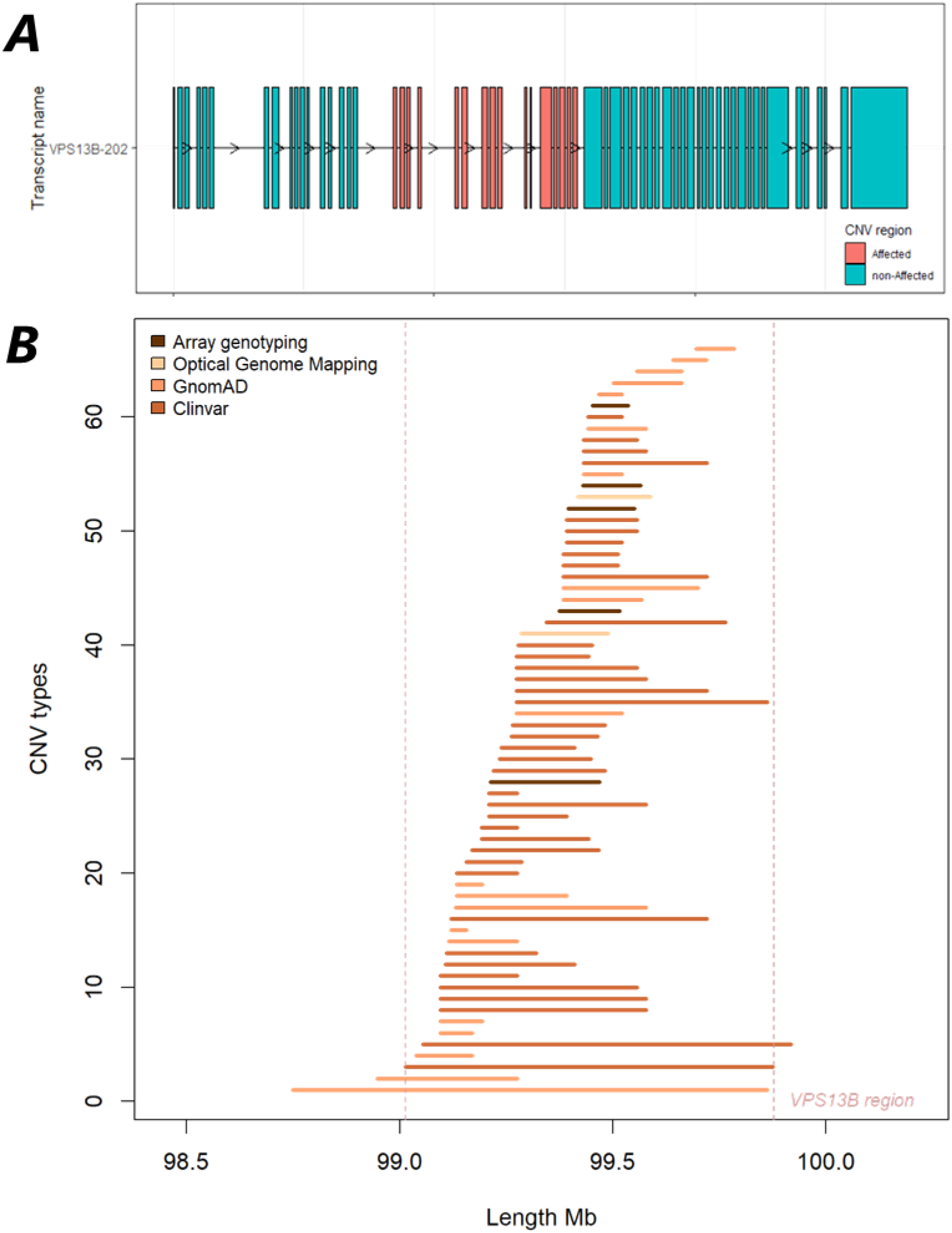
*VPS13B* gene region**. A.** Mane transcript visualization (VPS13B-202 or ENST00000357162; 62 exons). Region affected by the CNVs found in GR@ACE/DEGESCO cohort is highlighted in red (exons 18-33). **B.** Different deletions detected in the *VPS13B* gene region (genomic positions in GRCh38/hg38 assembly). Array genotyping and optical genome mapping corresponds to the GR@ACE/DEGESCO individuals while Clinvar and GnomAD to open databases.

**Table 2:** Dementia patients with deletions in *VPS13B* gene found by genotyping array and/or optical genome mapping technology (OGM). Positions were in genome assembly GRCh38/hg38. FTD: FrontoTemporal Dementia, AD: Alzheimer’s Disease.

| Indiv. | CNV Start | CNV End | Nsnps | Length (kb) | Technique | Sex | Phenotype |
| --- | --- | --- | --- | --- | --- | --- | --- |
| 1 | chr8:99466383 | 99586493 | 16 | 120.1 | Array | Male | FTD |
| 1 | chr8:99418180 | 99598117 | -- | 177.9 | OGM | Male | FTD |
| 2 | chr8:99429591 | 99564413 | 20 | 134.8 | Array | Female | AD |
| 3 | chr8:99452443 | 99586493 | 17 | 134.1 | Array | Male | AD |
| 3 | chr8:99418180 | 99598117 | -- | 177.9 | OGM | Male | AD |
| 4 | chr8:99374974 | 99515703 | 22 | 140.7 | Array | Male | AD |
| 5 | chr8:99429591 | 99564413 | 20 | 134.8 | Array | Male | AD |
| 6 | chr8:99452443 | 99535165 | 10 | 82.7 | Array | Female | AD |
| 7 | chr8:99212992 | 99467786 | 32 | 254.8 | Array | Male | AD |
| 8 | chr8:99395405 | 99550235 | 22 | 154.8 | Array | Female | AD |
| 9 | chr8:99338516 | 99467786 | 20 | 129.3 | Array | Female | AD |
| 9 | chr8:99285332 | 99489787 | -- | 196.0 | OGM | Female | AD |

OGM enabled detailed molecular characterization of the CNVs in the *VPS13B* gene. Direct visualization of molecules revealed a rearrangement in this genomic region in all samples tested (n=3, Figure 3). In all three samples, the region identified by the array is fully contained within the larger region detected by OGM. The latter expands the initial region by ∼44**%**, suggesting a broader or more sensitive capture of the associated signal. In the case of individuals 1 and 3, we observed a 177.9 Kb deletion on chr8:99418180-99589117 encompassing exons 22-33 from *VPS13B* gene. On the other hand, for individual 9, the deletion covered a region of 196Kb (chr8:99285332-99489787) and affected exons 20-25 (Table 2). We therefore sought to further investigate the mutations of this gene.

**Figure 3:**
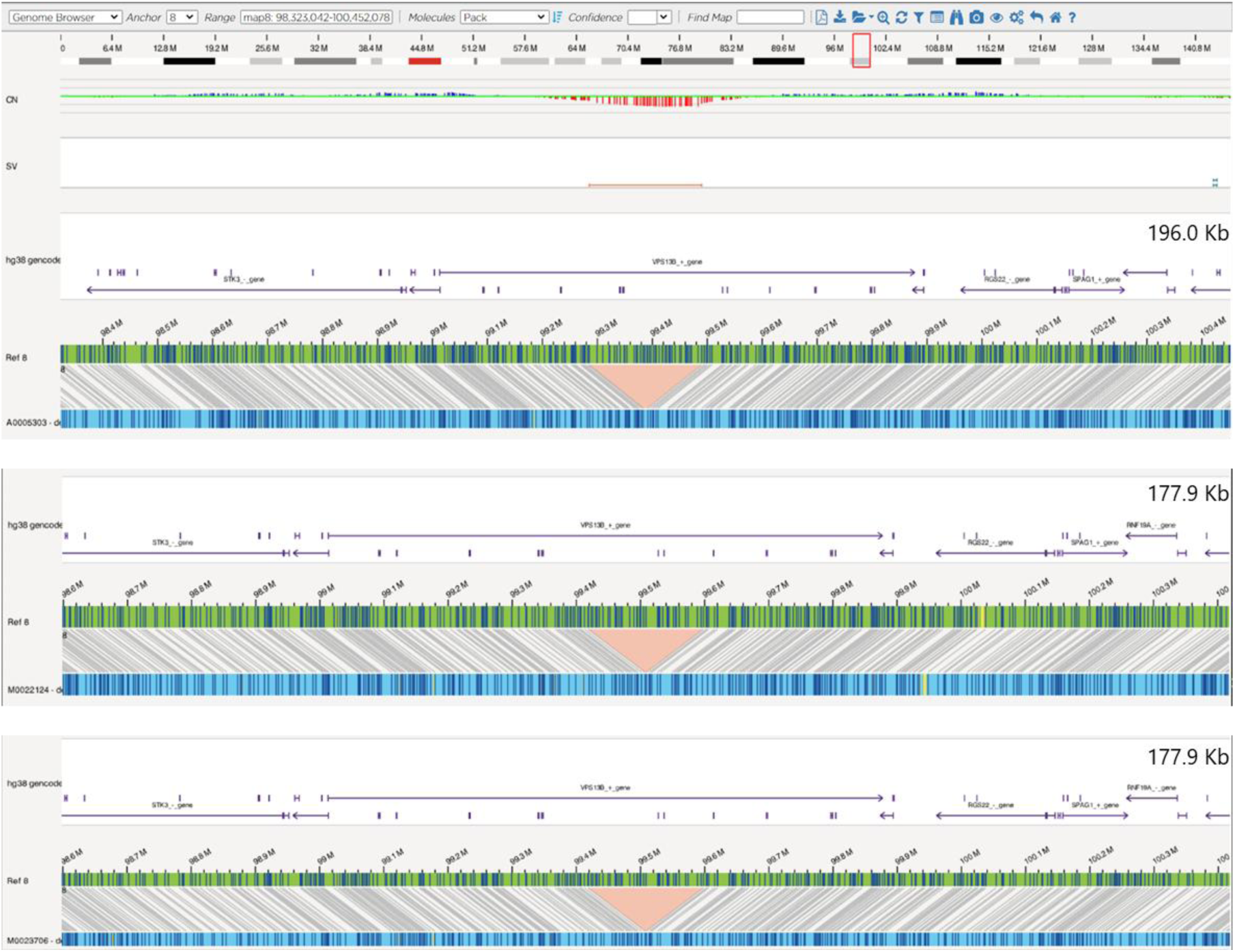
*VPS13B* CNV regions of three individuals from GR@ACE/DEGESCO by Optical Genome Mapping. Genomic positions in GRCh38/hg38 assembly.

In ClinVar, 3,681 entries were identified for the *VPS13B* gene, with 512 being CNVs, constituting 13.9% of the total. Among these CNVs, 63% were classified as pathogenic or likely pathogenic. Specifically, 76% of the CNVs were linked to Cohen Syndrome - rare autosomal recessive disorder caused by biallelic loss-of-function variants in *VPS13B* - while the majority of the remaining 24% comprised unspecified cases and other pathologies. Approximately, 10% of the CNVs (n=51) overlapped with the region affected in our individuals, with 43 (84.3%) deletions, of which 32 (74%) were referred to the condition of Cohen syndrome (Supplementary Table 7). Furthermore, it seems that all deletions we have detected bear striking similarity to those previously described as uncertain, pathogenic or likely pathogenic in ClinVar or GnomAD (Figure 2). In addition, 668 SNPs were found in the TOPMed imputation reference panel, with a minor allele frequency (MAF) <1%, classified as uncertain, pathogenic or likely pathogenic, from which only 169 of these variants met the high quality (Rsq>0.3). We found two SNPs significantly associated with Cohen’s Syndrome (conflicting pathogenicity) for AD (rs141637316(G), OR=3.85[1.01-14.65], p=0.0487; and rs142248228(A), OR=3.93[1.01-15.29], p=0.0499).

Although Poisson regression analysis suggested a nominally significant increase in variant counts in AD cases (p=1.52×10^-03^), this effect was not significant using models accounting for overdispersion (overdispersion ratio = 6.83; NBR OR=1.02[0.98-1.05], p=0.334) or non-parametric testing (p=0.282), suggesting limited burden difference between groups (Supplementary Figure 2).

### Candidate gene approach (AD genes)

We identified 45 of the 76 candidate genes tested (59%) overlapped by CNV calls in the GR@ACE/DEGESCO study (Supplementary Table 8). Global AD burden analyses did not reveal significant differences between dementia cases and controls in CNV rate, distribution of deletions or duplications (deletions Fisher test p=0.0875, duplications Fisher Test p=0.861). However, we identified a deletions affecting the *MAPT* gene in 14 individuals, including 13 controls and one dementia case (Freq_ctrl-dem_ = 0.16-0.01%, Supplementary Figure 3). Notably, all carriers shared the H2 haplotype (H1=49.4%, H1/H2=41.7%, H2=8.85% in GR@ACE/DEGESCO control population), which has been associated with reduced MAPT expression and decreased risk for tauopathies^56^. In contrast, risk-associated *ABCA7* deletions (4 controls and 11 cases, Freq_ctrl-dem_ = 0.05-0.14%, p = 4.7×10^-03^) and *APP* CNVs where observed (3 dementia cases, Freq_dem_ = 0.038%; deletions=2, duplication=1, Supplementary Table 9).

### ClassifyCNV

A total of 19,734 unique CNVs were identified in AD and control individuals (54.2% deletions). Among these, only 269 CNVs (1.36%) were classified as pathogenic or likely pathogenic, while the majority, 17,517 CNVs (88.8%), were of uncertain significance.

The untargeted analysis of pathogenic deletions (n=243, 90.3%), based on the full CNV region, did not yield any statistically significant association. The strongest signal was observed in the *MAPT*-*STH*-*KANSL1* locus (chr17:44071294:44358286, GRCh37/hg19), with three occurrences in control subjects and none in AD cases (Fisher’s p=0.115). Similarly, the sensitive gene-based approach identified four genes with nominal Fisher’s p-values <0.05 (*KANSL1*, *COL5A1*, *NOTCH1* and *PKD1*), although none remained significant after Bonferroni correction.

## Discussion

CNVs have emerged as relevant contributors to neurodegenerative disorders, yet their role in dementia remains uncertain. Some studies have investigated CNVs and other structural variants in different forms of dementias^6,8,20,29,30,57–62^, but their pathogenicity is often difficult to establish, particularly in the absence of segregation evidence, making it unclear whether they act as primary drivers or modifiers of disease risk. While CNVs involving well-known disease-associated genes can be linked more readily to disease mechanism, interpreting the significance of CNVs in genes without established disease-causing mutations remains particularly challenging.

In this study, by analyzing CNVs in a large Spanish cohort of array-genotyped individuals with detailed clinical and diagnostic information, enabled us to identify potentially novel CNV-affected genes associated with dementia and confirm CNVs as a source of genetic risk for dementia. Differences in CNV burden across studies—including ADNI^16^, NIA-LOAD/NCRAD^63^, TGen^8^ and Alzheimer’s Disease Sequencing Project^62^ —likely reflect variation in study design, genotyping platforms, QC procedures, or cohort composition, underscoring the need for harmonized analytical strategies^64^.

Our candidate AD-gene approach revealed CNVs in *APP* and *ABCA7* genes, supporting potential role of these CNVs in the pathogenesis of AD. In contrast, partial and total deletions affecting the *MAPT* locus were observed almost exclusively in controls, and all carriers harbored the H2 haplotype. The H2 haplotype is known to contain a large inversion^65,66^ (∼900 kb) and has been linked to reduced risk for several tau-related disorders^56,67–72^, including AD^73–75^. Structural variation on the H2 background is less frequent than on H1^76^, yet the presence of deletions in this context may reflect a stable, inherited structural variant with a potential protective effects. Alternatively, it could point to a regional *MAPT* effect, given that the H2 haplotype is more common in Spanish population^77^. Since *MAPT* dosage and expression levels are tightly linked to tau pathology, it is plausible that reduced copy number in the context of H2 might contribute to resilience against neurodegeneration. Additional CNVs in the *MAPT–STH–KANSL1* region were also highlighted by the classifyCNV approach, a tool for prioritizing potential pathogenicity. However, these variants appeared more frequently in controls and did not reach statistical significance, suggesting they may represent benign structural variation with context- specific effects unrelated to AD or limited functional relevance. Functional studies will be required to explore this hypothesis further, the consequences of these deletions and its potential role in modulating disease susceptibility, gene expression and disease progression.

Interestingly, two of our top candidate genes in the genome-wide approach, *PKP3*^78^ and *FBRSL1*^79^, have potential functional connections to neurodevelopmental delay and related conditions such as ADRD. *PKP3* (Plakophilin 3) is a gene encoding a protein involved in cell adhesion, particularly in maintaining the structural integrity of tissues^80,81^. It’s primarily associated with diseases such as arrhythmogenic right ventricular cardiomyopathy and certain skin disorders^82^. Its related pathways include the nervous system development. On the other hand, *FBRSL1* (Fibrinogen Receptor Like 1) encodes a integrin-related protein involved in cell adhesion and signaling. Pathogenic variants in this gene have been associated with an intellectual disability syndrome characterized by developmental delay, autistic traits, and multiple systemic features^83–87^. Furthermore, expression data from GTEx and cloning studies indicate moderate to high FBRSL1 expression across adult brain regions, with highest levels in cerebellum and hippocampus^79,88^.

Our third candidate, *SIGIRR* (also known as IL-1R8 or TIR8), is a negative regulator of innate immune signaling expressed in neurons, astrocytes and microglia^89–91^. It inhibits TLR and IL-1R pathways^92^, thereby attenuating IL-1β-mediated inflammation. In mice, SIGIRR deficiency leads to increased IL-1β signaling, impaired synaptic plasticity, and cognitive deficits, highlighting its potential role in modulating neuroinflammation and cognition^93^. Given the chronic upregulation of IL-1β and related cytokines in AD, the SIGIRR’s involvement in the anti-inflammatory IL-37 pathway, and the IL-1β-IL-1R axis has been implicated in causing tau pathology in AD, loss-of-function or deletion of *SIGIRR* could exacerbate neuroinflammation and contribute to AD pathogenesis^94,95^.

Our study identifies genes enriched in cases relative to controls, suggesting the possible involvement of these genes in AD and/or dementia susceptibility. While the small number of carriers limits statistical power, the absence of this CNVs in controls raises the possibility of a potential disease-specific effect. For example, the duplication in the 14q11.2 region enriched in cases, encompassing multiple genes including *IL25*, a regulator of neuroinflammatory responses in neurodegenerative diseases^96^. Elevated IL25 levels have been associated with cognitive decline, indicating its potential as a biomarker for disease progression^97^. Given the CNV also spans other genes (∼79 to 146 Kb), the biological and clinical relevance of this duplication likely extends beyond *IL25* alone and needs replication. This raises the possibility of a contiguous gene effect, where the combined dosage changes of multiple neighboring genes may contribute to the phenotype. Functional characterization of all affected genes will be necessary to clarify the biological relevance and pathogenic potential of this duplication.

Notably, this the first report describing intragenic deletions in *VPS13B* among ADRD cases (n=9, Freq_cases_= 0.115%), validated by an independent technique (OGM). These deletions span exons 18–33 out of 62, removing a substantial portion of the coding region and likely disrupt protein structure and function—either by causing a frameshift and premature termination (leading to nonsense-mediated decay) or by deleting critical domains involved in vesicular trafficking and Golgi organization.

Loss-of-function mutations in *VPS13B* are known to cause Cohen syndrome^98,99^, characterized by phenotypic consequences including developmental delay, intellectual disability, microcephaly, hypotonia and systemic features, which supports a potential deleterious effect of these variants on neuronal homeostasis, although their relevance to dementia requires further investigation. While the GnomAD CNV database lists 20 types of deletions affecting *VPS13B* gene region (n=108 in 464,297 individuals, Freq=0.023%, Figure 2), no previous experimental or genetic studies have linked this locus to ADRD. However, additional evidence emerges from two rare pathogenic SNPs in *VPS13B* (rs141637316 and rs142248228) previously linked to Cohen syndrome, which also showed significant associations with AD. The convergence of structural and rare variant signals, together with reported associations with AD-related endophenotypes such as hippocampal volume^100^, further support its candidacy as a dementia susceptibility gene.

Strikingly, mutations in other VPS13 family members are also implicated in neurological disorders, including chorea-acanthocytosis (VPS13A)^101^, syndromic autism^102–104^ (VPS13B), early onset Parkinson’s disease (VPS13C)^105,106^ and ataxia (VPS13D)^107^. This diversity highlights the broad role of VPS13 proteins in brain health. *VPS13B* encodes a large transmembrane protein likely involved in vesicle-mediated transport and protein sorting^108^. Disruptions in lipid transport at membrane contact sites—where VPS13 proteins operate— are linked to several neurological disorders, emphasizing the importance of organelle lipid homeostasis for neuronal function^109,110^. Additionally, *VPS13B* plays a role in nervous system development and may be essential for neuron projection growth^111,112^.

Furthermore, pathway analysis in the duplications exclusively in dementia cases revealed the presence of well-established roles in cardiac and vascular biology, suggesting a potential link to vascular co-pathology^32,113^ in AD. These findings may reflect a shared genetic vulnerability affecting both cardiac and cerebral vascular systems, potentially contributing to blood-brain barrier breakdown, neuroinflammation, and impaired perfusion, all of which are common in patients with mixed or vascular-influenced dementia^114,115^.

Despite the progress in understanding CNVs in dementia, significant challenges remain for clinical translation. The heterogeneity of CNVs and their interaction with genetic, epigenetic, and environmental factors complicate the definition of clear mechanistic hypotheses, underscoring the need for integrative and multidisciplinary approaches. We also acknowledge limitations of array-based CNV detection. While arrays provide a cost-effective genotyping of large cohorts, their resolution depends on marker density^116^, leading to conservative estimates and reduced sensitivity in poorly covered regions, such as near centromeres. Moreover, arrays cannot resolve breakpoints at base-pair resolution or differentiate structural events like inversions or translocations, and detection of small CNVs (<1 kb) remains particularly challenging due to signal variability.

## Conclusions

The present work represents the first CNV analysis within the GR@ACE/DEGESCO cohort, the largest clinically characterized dementia cohort in Spain. We identified deletions in *VPS13B, PKP3*, *FBRSL1* and *SIGIRR* genes and duplications in 14q11.2 region, suggesting possible contributions to ADRD, emphasizing the importance of further research to elucidate their underlying mechanisms. As evidence linking CNVs and ADRD grows, genotyping array data remains valuable for identifying structural variants with potential pathogenic roles. Nonetheless, complementary strategies such as long-read sequencing, functional characterization, and standardized meta-analyses will be essential to replicate findings and fully leveraging the potential of structural genomics to improve diagnosis, prognosis, and treatment of dementia, thus offering hope for better outcomes for people affected by these complex neurodegenerative conditions.

## Supporting information

SupplementaryData_CNVs_GRACE_DEGESCO

## Data Availability

The data that support the findings of this study are available on request from the GR@ACE/DEGESCO consortium. Summary statistics of the CNVs by gene association analysis will be publicly available at Zenodo after publication.

## List of abbreviations

ACMG: American College of Medical Genetics and Genomics classification guidelines
AD: Alzheimer’s disease
ADNI: Alzheimer’s Disease Neuroimaging Initiative
ADRD: Alzheimer’s Disease and Related Dementias
ANOVA: Analysis of variance
BAF: B Allele Frequency
CeGEN: Spanish National Center for Genotyping
CGSs: Contiguous gene syndromes or CNV syndromes
CNVs: Copy number variations
DEGESCO: Dementia Genetic Spanish Consortium
DLE-1: Direct Labeling Enzyme 1
DLS: Direct label and stain
DN: de novo
DSM-IV: Diagnostic and Statistical Manual of Mental Disorders, Fourth Edition criteria
FBRSL1: Fibrinogen Receptor Like 1
FTD: Frontotemporal Dementia
GLMs: Generalized linear models
GnomAD: Genome Aggregation Database SV resource
GO: Gene Ontology
GR@ACE: Genome Research at Ace Alzheimer Center Barcelona
GTEx: Genotype-Tissue Expression project
GWAS: Genome-Wide Association Studies
HGNC: HUGO Gene Nomenclature Committee
kb: Kilobase
KEGG: Kyoto Encyclopedia of Genes and Genomes
LRR: Log R Ratio
MAF: Minor allele frequency
Mb: Megabase
MCI: Mild cognitive impairment
NBR: Negative binomial regressions
NGS: Next-generation sequencing
NIA–AA: National Institute on Aging and Alzheimer’s Association
NIA-LOAD/NCRAD: National Institute on Aging – Late-Onset Alzheimer’s Disease / National Cell Repository for Alzheimer’s Disease
OGM: Optical genome mapping
PCA: Principal component analysis
PFB: Population frequency of B allele
PKP3: Plakophilin 3
PMSF: Phenylmethylsulfonyl fluoride treatment
QC: Quality control
Rsq: Imputation quality score
SD: Standard deviation
SNP: Single nucleotide polymorphism
SV: Structural variations
TGen: Translational Genomics Research Institute
TOPMed: Trans-Omics for Precision Medicine program
UCSC: University of California Santa Cruz Genome Browser
UHMW: Ultra-high molecular weight
VPS13B: Vacuolar Protein Sorting 13 Homolog B gene
WF: Waviness Factor

## Declarations

### Ethics approval and consent to participate

The ethics and scientific committees have approved, this research protocol (Acta 25/2016, Ethics Committee H., Clinic I Provincial, Barcelona, Spain). Informed consent was obtained from all subjects involved in the study.

### Consent for publication

All participants provided written informed consent for the use and publication of anonymized data as part of the study protocol.

### Availability of data and materials

The data that support the findings of this study are available on request from the GR@ACE/DEGESCO consortium. Summary statistics of the CNVs by gene association analysis are publicly available at Zenodo (doi).

### Authors’ contributions

AR, SvdL and IdR designed and conceptualized the study. AR, MVF and SvdL supervised the study, interpreted the data and revised the manuscript. IdR contributed to data acquisition, the analysis, interpreted the data, prepared figures and wrote the manuscript. The rest of the authors contributed to the data acquisition and investigation. All authors critically revised the manuscript for important intellectual content and approved the final manuscript.

## Acknowledgements

We would like to thank all participants who collaborated in this project. Samples were gathered as part of the Genome Research at Ace (GR@ACE), Dementia Genetics Spanish Consortium (DEGESCO), and the National DNA Bank Carlos III (www.bancoadn.org, University of Salamanca, Spain). All samples were processed following standard operating procedures with the appropriate approval of the Ethical and Scientific Committee. Ace Alzheimer Center Barcelona is one of the participating centers of the Dementia Genetics Spanish Consortium (DEGESCO).

## The GR@ACE study group

Aguilera Nuria^1^, Alegret Montserrat^1,2^, Bayón-Buján Paula^1^, Bein Natali^1^, Blazquez-Folch Josep^1^, Boada Mercè^1,2^, Buendia Mar^1^, Cano Amanda^1^, Calm Berta^1^, Cañabate Pilar^1,2^, Capdevila-Bayo Maria^1,3^,Carracedo Angel^4,5^, Corbatón-Anchuelo A^6^, Casales Federico^1^, de Rojas Itziar^1,2,7^, Diego Susana^1^, Espinosa Ana^1,2^, Fernandez Maria Victoria^1^, Gailhajenet Anna^1^, García-González Pablo^1,3^, Guitart Marina^1^, Ibarria Marta^1^, Lafuente Asunción^1^, Lleonart Núria^1^, Macias Juan^8^, Maroñas Olalla^4^, Martín Elvira^1^, Martínez Maria Teresa^6^, Marquié Marta^1,2^, Miguel Andrea^1^, Montrreal Laura^1^, Morató Xavier^1,2^, Moreno Mariola^1^, Muñoz Nathalia^1^, Muñoz-Morales Álvaro^1^, Olivé Clàudia^1,3^, Ortega Gemma^1,2^, Pancho Ana^1^, Pelejà Ester^1^, Pérez-Cordon Alba^1^, Pineda Juan A^8^, Puerta Raquel^1,3^, Preckler Silvia^1^, Quintela Inés^4^, Real Luis Miguel^8^, Rodríguez José Nelet^1^, Rosende-Roca Maitee^1,2^, Ruiz Agustín^1,2,7^, Sáez Maria Eugenia^10^, Sanabria Angela^1,2^, Seguer Susanna^1^, Sotolongo-Grau Oscar^1^, Tárraga Luís^1,2^, Tartari Juan Pablo^1^, Valero Sergi^1,2^, Vargas Liliana^1^.

^1^Ace Alzheimer Center Barcelona, Universitat Internacional de Catalunya (UIC), Barcelona, Spain. ^2^Networking Research Center on Neurodegenerative Diseases (CIBERNED), Instituto de Salud Carlos III, Madrid, Spain. ^3^Doctorate in Biotecnology, Faculty of Pharmacy and Food Sciences, University of Barcelona, Barcelona, Spain. ^4^Grupo de Medicina Xenómica, Centro Nacional de Genotipado (CEGEN-PRB3-ISCIII). Universidad de Santiago de Compostela, Santiago de Compostela, Spain. ^5^Fundación Pública Galega de Medicina Xenómica- CIBERER-IDIS, Santiago de Compostela, Spain. ^6^Centro de Investigación Biomédica en Red de Diabetes y Enfermedades Metabólicas Asociadas, CIBERDEM, Spain, Hospital Clínico San Carlos, Madrid, Spain. ^7^Luxembourg Centre for Systems Biomedicine (LCSB), University of Luxembourg, Esch-sur-Alzette, Luxembourg. ^8^Unidad Clínica de Enfermedades Infecciosas y Microbiología. Hospital Universitario de Valme, Sevilla, Spain. ^9^Biggs Institute for Alzheimer’s and Neurodegenerative Diseases, University of Texas Health Science Center, San Antonio, Texas, USA. ^10^CAEBI. Centro Andaluz de Estudios Bioinformáticos, Sevilla, Spain.

## DEGESCO consortium

Adarmes-Gómez Astrid Daniela^1,2^, Aguilar Miquel^3,4^, Aguilera Nuria^5^, Alcolea Daniel^6,2^, Alegret Montserrat^7,2^, Alonso María Dolores^8^, Alvarez Ignacio^3,4^, Álvarez Victoria^9,10^, Amer-Ferrer Guillermo^11^, Antequera Martirio^12^, Antonell Anna^13^, Antúnez Carmen^14^, Arias Pastor Alfonso^15,16^, Baquero Miquel^17,18^, Bayón-Buján Paula^7^, Belbin Olivia^6,2^, Bernal Sánchez-Arjona María^19^, Boada Mercè^7,2^, Bonilla-Toribio Marta^1,2^, Buendia Mar^5^, Buiza-Rueda Dolores^20,2^, Bullido María Jesús^21,2,22,23^, Buongiorno Maria Teresa^3,4^, Calero Miguel^24,2,25^, Camara Ana^26,2^, Cañabate Pilar^5^, Cano Amanda^7^, Cardona Serrate Fernando^27,2,28^, Carracedo Ángel^29,30^, Carrillo Fátima^1,2^, Casajeros María José^31^, Chafer Pericás Consuselo^17^, Compta Yaroslau^26^, Corbatón-Anchuelo Arturo^32,33^, Corma-Gómez Anaïs^34^, De la Guía Paz^35^, de Rojas Itziar^7,2,36^, del Ser Teodoro^37^, Díaz Belloso Rafael^1,2^, Diego Susana^5^, Diez-Fairen Mónica^3,4^, Dols-Icardo Oriol^6,2^, Erro Aguirre María Elena^38,39,40^, Espinosa Ana^5,2^, Fernandez Manuel^26,2,41^, Fernández Maria Victoria^7^, Fernández-Fuertes Marta^34^, Fortea Juan^6,2^, Franco-Macías Emilio^19,2^, Frank-García Ana^42,2,43,23^, Gailhajenet Anna^5^, García-Alberca Jose María^35^, García-Díaz Sergio^1,2^, García-González Pablo^7,44^, García-Madrona Sebastián^31^, Garcia-Ribas Guillermo^31^, García-Roldán Ernesto^45^, Garrote-Espina Lorena^1,2^, Gómez-Garre Pilar^1,2^, Guitart Marina^5^, Huerto Vilas Raquel^15,16^, Ibarria Marta^5^, Illán-Gala Ignacio^46,2^, Jesús Silvia^1,2^, Kulisevsky Jaime^47,2^, Labrador Espinosa Miguel Angel^1,2^, Lafuente Asunción^5^, Lage Carmen^48,2^, Legaz Agustina^12^, Lleó Alberto^6,2^, López-García Sara^49,2^, Lopez de Munain Adolfo^50,51,2,52^, Macías Juan^34^, Macias-García Daniel^1,2^, Manzanares Salvadora^12^, Marín Marta^45^, Marín-Muñoz Juan^12^, Maroñas Olalla^53^, Marquié Marta^7,2^, Marti Maria^26^, Martín Elvira^5^, Martín-Bórnez Miguel^1,2^, Martín-Montes Angel^54,2,43^, Martín Montés Angel^54,2,43^, Martínez Victoriana^12^, Martínez Begoña^12^, Martínez-Lage Álvarez Pablo^55^, Martínez-Larrad María Teresa^32,33^, Martinez de Pancorbo Marian^56^, Martínez Rodríguez Carmen^57,10^, Medina Miguel^2,25^, Mendioroz Iriarte Maite^38,39,40^, Mendoza Silvia^35^, Menéndez-González Manuel^58,10,59^, Mir Pablo^1,2,60^, Molina-Porcel Laura^61,13^, Montrreal Laura^7^, Moreno Mariola^5^, Moreno Fermin^50,2,52^, Muñoz Esteban^26^, Muñoz-Delgado Laura^1,2^, Noguera Perea Fuensanta^12^, Olivé Clàudia^7,44^, Ortega Gemma^5,2^, Pagonabarraga Javier^47,2^, Painous Celia^26^, Pancho Ana^5^, Pastor Ana Belén^62,63^, Pastor Pau^64,65^, Pelejá Ester^5^, Pérez-Cordón Alba^5^, Pérez-Tur Jordi^27,2^, Periñán María Teresa^1,2,66^, Pineda Juan Antonio^34^, Pineda-Sánchez Rocío^1,2^, Piñol-Ripoll Gerard^15,16^, Preckler Silvia^5^, Puerta Raquel^7,44^, Quintela Inés^67^, Rábano Alberto^62,63,2^, Real Luis Miguel^68,69^, Real de Asúa Diego^70^, Rodriguez-Rodriguez Eloy^48,2^, Rosas Allende Irene^9,10^, Rosende-Roca Maitée^5,2^, Royo Jose Luis^71^, Ruiz Agustín^7,2,72^, Sáez María Eugenia^73^, Sanabria Ángela^5,2^, Sánchez-Juan Pascual^74,2^, Sánchez-Valle Raquel^13^, Sánchez Ruiz de Gordoa Javier^38^, Sastre Isabel^21,2^, Sotolongo-Grau Oscar^7^, Tárraga Lluís^7,2^, Valero Sergi^7,2^, Valldeoriola Francesc^26^, Vargas Liliana^5^, Vicente María Pilar^12^, Vivancos-Moreau Laura^12^.

^1^Unidad de Trastornos del Movimiento, Servicio de Neurología, Instituto de Biomedicina de Sevilla, Hospital Universitario Virgen del Rocío/CSIC/Universidad de Sevilla, Seville, Spain, ^2^CIBERNED, Network Center for Biomedical Research in Neurodegenerative Diseases, National Institute of Health Carlos III, Madrid, Spain, ^3^Fundació Docència i Recerca MútuaTerrassa, Terrassa, Barcelona, Spain, ^4^Memory Disorders Unit, Department of Neurology, Hospital Universitari Mutua de Terrassa, Terrassa, Barcelona, Spain, ^5^Ace Alzheimer Center Barcelona, Universitat Internacional de Catalunya (UIC), Barcelona, Spain., ^6^Department of Neurology, II B Sant Pau, Hospital de la Santa Creu i Sant Pau, Universitat Autònoma de Barcelona, Barcelona, Spain., ^7^Ace Alzheimer Center Barcelona, Universitat Internacional de Catalunya, Barcelona, Spain., ^8^Servei de Neurologia. Hospital Clínic Universitari de València, ^9^Laboratorio de Genética. Hospital Universitario Central de Asturias, Oviedo, Spain, ^10^Instituto de Investigación Sanitaria del Principado de Asturias (ISPA), ^11^Department of Neurology, Hospital Universitario Son Espases, Palma, Spain, ^12^Unidad de Demencias. Hospital Clínico Universitario Virgen de la Arrixaca, Palma, Spain, ^13^Alzheimer’s disease and other cognitive disorders unit. Service of Neurology. Hospital Clínic of Barcelona. Institut d’Investigacions Biomèdiques August Pi i Sunyer, University of Barcelona, Barcelona, Spain, ^14^Unidad de Demencias, Hospital Clínico Universitario Virgen de la Arrixaca, Murcia, Spain., ^15^Unitat Trastorns Cognitius, Hospital Universitari Santa Maria de Lleida, Lleida, Spain, ^16^Institut de Recerca Biomedica de Lleida (IRBLLeida), Lleida, Spain, ^17^Servei de Neurologia, Hospital Universitari i Politècnic La Fe, Valencia, Spain, ^18^Grupo de Investigación en Enfermedad de Alzheimer, GINEA, Instituto de Investigación Sanitaria La Fe, ^19^Unidad de Demencias, Servicio de Neurología y Neurofisiología. Instituto de Biomedicina de Sevilla (IBiS), Hospital Universitario Virgen del Rocío/CSIC/Universidad de Sevilla, Seville, Spain, ^20^Unidad de Trastornos del Movimiento, Servicio de Neurología y Neurofisiología. Instituto de Biomedicina de Sevilla (IBiS), Hospital Universitario Virgen del Rocío/CSIC/Universidad de Sevilla, Seville, Spain, ^21^Centro de Biología Molecular Severo Ochoa (UAM-CSIC), ^22^Instituto de Investigacion Sanitaria ‘Hospital la Paz’ (IdIPaz), Madrid, Spain, ^23^Universidad Autónoma de Madrid, ^24^CIEN Foundation/Queen Sofia Foundation Alzheimer Center/Instituto de Salud Carlos III, ^25^UFIEC, Instituto de Salud Carlos III, ^26^Parkinson’s Disease & Movement Disorders Unit, Neurology Service, Hospital Clínic I Universitari de Barcelona; IDIBAPS, CIBERNED (CB06/05/0018-ISCIII), ERN- RND, Institut Clínic de Neurociències (Maria de Maeztu Excellence Centre), Universitat de Barcelona. Barcelona, Catalonia, Spain, ^27^Unitat de Genètica Molecular, Institut de Biomedicina de València-CSIC, Valencia, Spain, ^28^Unidad Mixta de Neurologia Genètica, Instituto de Investigación Sanitaria La Fe, Valencia, Spain., ^29^Grupo de Medicina Xenómica, CIBERER, CIMUS. Universidade de Santiago de Compostela, Santiago de Compostela, Spain., ^30^Fundación Pública Galega de Medicina Xenómica- IDIS, Santiago de Compostela, Spain., ^31^Hospital Universitario Ramon y Cajal, IRYCIS, Madrid, ^32^Instituto de Investigación Sanitaria, Hospital Clínico San Carlos (IdISSC), Madrid, Spain, ^33^Spanish Biomedical Research Centre in Diabetes and Associated Metabolic Disorders(CIBERDEM), Madrid, Spain, ^34^Unidad Clínica de Enfermedades Infecciosas y Microbiología. Hospital Universitario de Valme, Sevilla, Spain, ^35^Alzheimer Research Center & Memory Clinic, Instituto Andaluz de Neurociencia, Málaga, Spain., ^36^Luxembourg Centre for Systems Biomedicine (LCSB), University of Luxembourg, Esch-sur-Alzette, Luxembourg, ^37^Department of Neurology/CIEN Foundation/Queen Sofia Foundation Alzheimer Center, ^38^Navarrabiomed, Pamplona, Spain, ^39^Department of Neurology, Hospital Universitario de Navarra, Pamplona, Spain, ^40^Instituto de Investigación Sanitaria de Navarra (IDISNA), Pamplona, Spain, ^41^Universitat de Barcelona (UB), ^42^Department of Neurology, La Paz University Hospital. Instituto de Investigación Sanitaria del Hospital Universitario La Paz. IdiPAZ., ^43^Hospital La Paz Institute for Health Research, IdiPAZ, Madrid, Spain, ^44^Doctorate in Biotecnology, Faculty of Pharmacy and Food Sciences, University of Barcelona, Barcelona, Spain., ^45^Unidad de Demencias, Servicio de Neurología. Instituto de Biomedicina de Sevilla (IBiS), Hospital Universitario Virgen del Rocío/CSIC/Universidad de Sevilla, Seville, Spain, ^46^Sant Pau Memory Unit, Department of Neurology, Institut de Recerca de Sant Pau; Hospital de Sant Pau, Barcelona, Spain, ^47^Movement Disorders Unit, Department of Neurology, Institut de Recerca de Sant Pau; Hospital de Sant Pau, Barcelona, Spain, ^48^Neurology Service, Marqués de Valdecilla University Hospital (University of Cantabria and IDIVAL), Santander, Spain., ^49^Service of Neurology, University Hospital Marqués de Valdecilla, IDIVAL, University of Cantabria, Santander, Spain, ^50^Department of Neurology. Hospital Universitario Donostia. San Sebastian, Spain, ^51^Department of Neurosciences. Faculty of Medicine and Nursery. University of the Basque Country, San Sebastián, Spain, ^52^Neurosciences Area. Instituto Biodonostia. San Sebastian, Spain, ^53^Grupo de Medicina Xenómica, Centro Nacional de Genotipado (CEGEN-PRB3-ISCIII). Universidade de Santiago de Compostela, Santiago de Compostela, Spain., ^54^Department of Neurology, La Paz University Hospital, ^55^Centro de Investigación y Terapias Avanzadas. Fundación CITA-alzheimer, San Sebastian, Spain, ^56^BIOMICS País Vasco; Centro de investigación Lascaray, Universidad del País Vasco UPV/EHU, Vitoria-Gasteiz, Spain, ^57^Hospital de Cabueñes, Gijón, Spain, ^58^Servicio de Neurología. Hospital Universitario Central de Asturias, Oviedo, Spain, ^59^Departamento de Medicina, Universidad de Oviedo, Oviedo, Spain, ^60^Departamento de Medicina, Facultad de Medicina, Universidad de Sevilla, Seville, Spain, ^61^Neurological Tissue Bank of the Biobanc-Hospital Clinic-IDIBAPS, Institut d’Investigacions Biomèdiques August Pi i Sunyer, Barcelona, Spain, ^62^CIEN Foundation/Queen Sofia Foundation Alzheimer Center, ^63^BT-CIEN, ^64^Unit of Neurodegenerative diseases, Department of Neurology, Hospital Germans Trias i Pujol, Badalona, Barcelona, ^65^Neurodegenerative Diseases Research Laboratory, Germans Trias i Pujol Research Laboratory, Badalona, Barcelona, ^66^Centre for Preventive Neurology, Wolfson Institute of Population Health, Queen Mary University of London, London, United Kingdom., ^67^Grupo de Medicina Xenómica, Fundación Pública Galega de Medicina Xenómica, Santiago de Compostela, Spain., ^68^Department of Medical Biochemistry, Molecular Biology and Immunology, University of Sevilla, Sevilla, Spain, ^69^CIBERINFEC, Network Center for Biomedical Research in Infectious Diseases, National Institute of Health Carlos III, Madrid, Spain, ^70^Hospital Universitario La Princesa, Madrid, Spain, ^71^Departamento de Especialidades Quirúrgicas, Bioquímica e Inmunología. School of Medicine. University of Malaga. Málaga, Spain, ^72^Biggs Institute for Alzheimer’s and Neurodegenerative Diseases, University of Texas Health Science Center, San Antonio, Texas, USA., ^73^CAEBI, Centro Andaluz de Estudios Bioinformáticos, Sevilla, Spain., ^74^Alzheimer’s Centre Reina Sofia-CIEN Foundation-ISCIII, Madrid, Spain

## Supplementary Figures

**Supplementary Figure 1:**
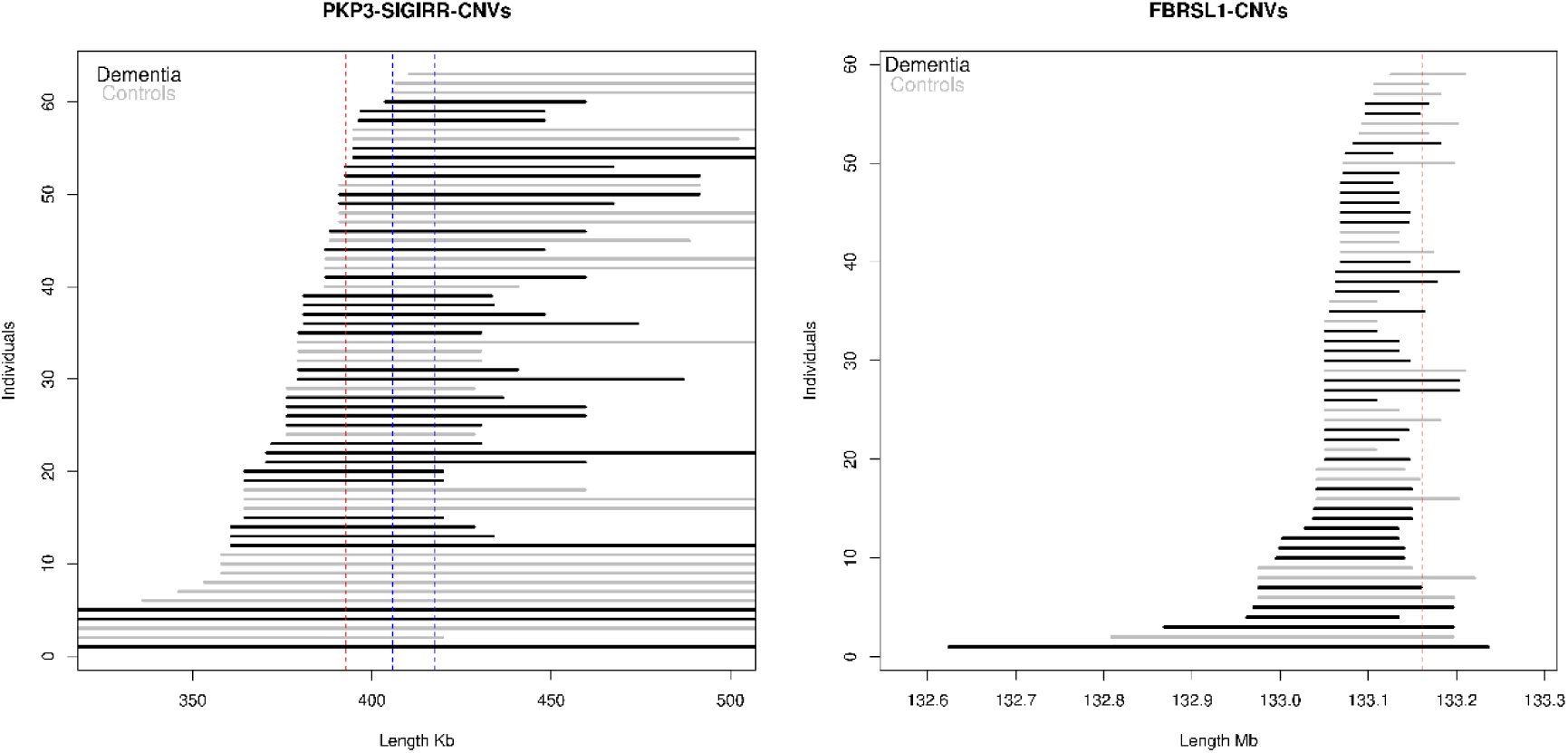
Copy number deletions affecting the top genes suggestively associated with dementia. *PKP3* and *FBRSL1* gene regions were highlighted in red, *SIGIRR* gene region in blue. Positions were in genome assembly GRCh37/hg19.

**Supplementary Figure 2:**
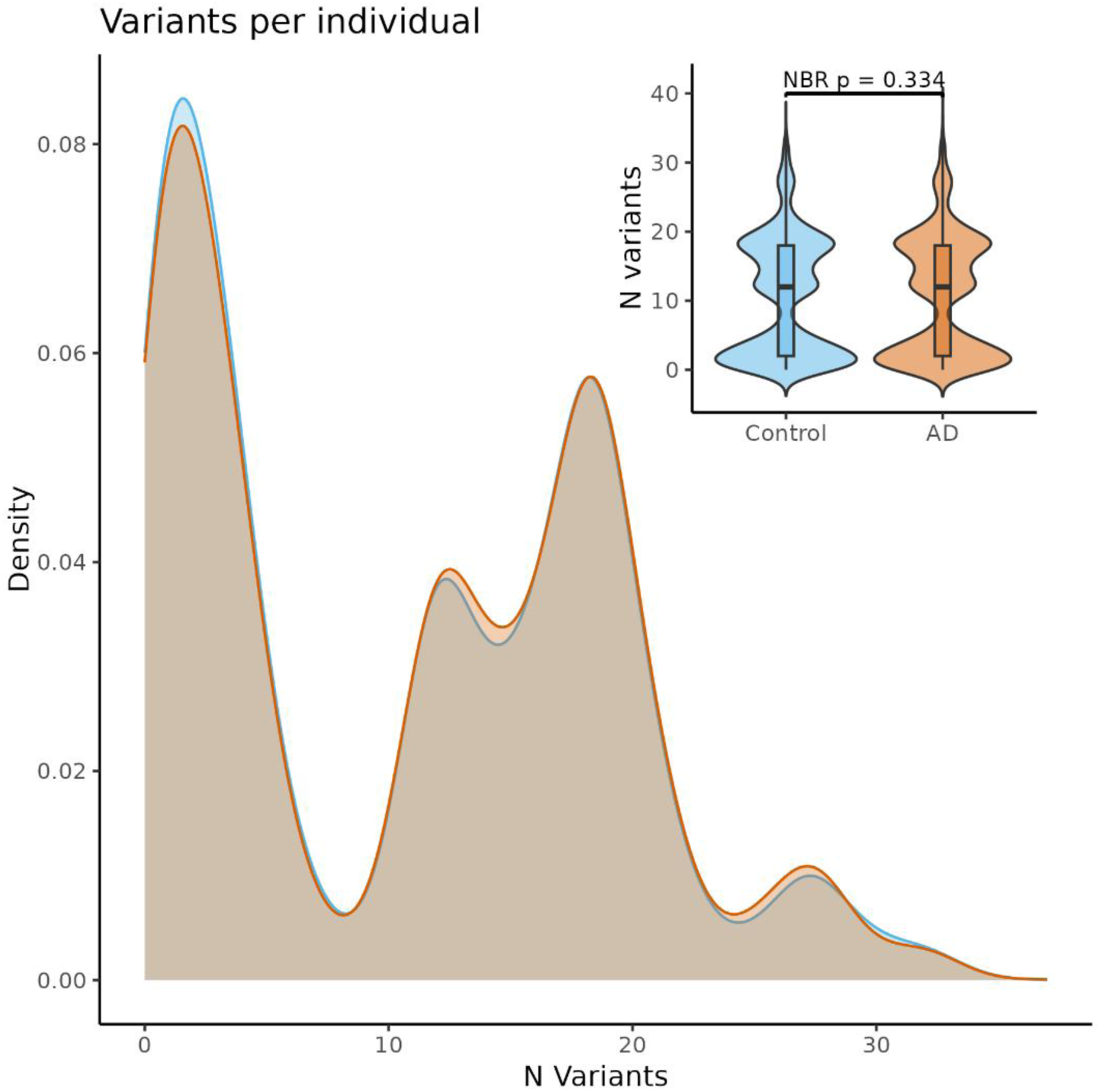
Distribution of variant burden per individual in the *VPS13B* gene region for the 169 SNPs significantly associated with Cohen’s Syndrome (ClinVar) available in the TOPMed imputation reference panel (Rsq>0.3, MAF<1%).

**Supplementary Figure 3:**
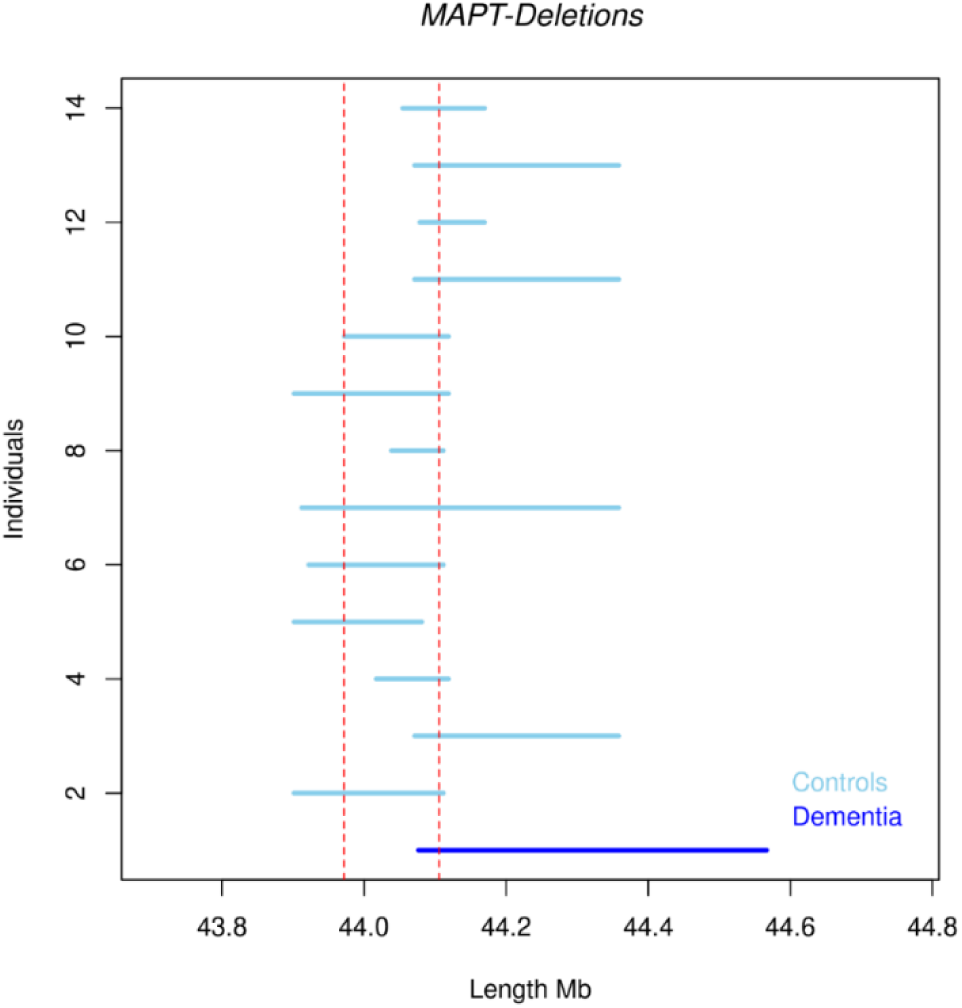
Copy number deletions affecting the *MAPT* genomic region. Gene region were highlighted in orange. Positions were in genome assembly GRCh37/hg19.

